# Prevalence of DSM-5 eating disorders and associated impairment and service use among children and young people in England: analysis of a general population survey

**DOI:** 10.64898/2026.10.20.26366038

**Authors:** Clara Faria, Aslihan Baser, Margarita Kousteni, Anya Ciobanca, Idura Hisham, Jessica O’Logbon, Simon White, Olurotimi Adejumo, Sally McManus, Tamsin Newlove-Delgado, Tamsin Ford

## Abstract

**Background:** Eating disorders (EDs) typically emerge in adolescence and carry substantial morbidity. DSM-5 revised EDs classification, introducing Other Specified Feeding or Eating Disorder (OSFED) and Avoidant Restrictive Food Intake Disorder (ARFID). We estimated general population prevalence of EDs, including ARFID, in children and young people (CYP), and the associated impairment, which remains poorly characterised.

**Methods:** We analysed the Mental Health of Children and Young People in England (MHCYP) 2023 survey, a nationally representative probability sample. All participants were screened for possible eating problems. Screen positives and a random sample of screen negatives were invited to a second-stage assessment; giving an analytic sample of 1,738 aged 11-25 years. Associations with impairment, mental health difficulties and service use were estimated using survey-weighted regression adjusted for age and sex.

**Results:** Any ED affected 6.7% (95% CI 5.4-8.3) of 11-25-year-olds and was over three times more common in females (10.7%) than males (3.0%). OSFED was the commonest (3.5%), followed by bulimia nervosa (1.7%), anorexia nervosa (0.9%), binge eating disorder (0.6%) and ARFID (0.2%). Prevalence peaked at 17-19 years (13.9%). EDs were associated with lifetime self-harm (aOR 9.1 at 11-16; 4.4 at 17-25), SDQ-predicted probable disorder (aOR 14.7; 2.5), medication use (aOR 5.3) and mental health service contact (aOR 2.1).

**Conclusions:** EDs are common among CYP in England and carry high levels of impairment, self-harm and comorbid mental health difficulties, particularly among 11-16-year-olds. Nonetheless, more than half of those meeting diagnostic criteria had no mental health service contact in the preceding year.

## Introduction

Eating Disorders (EDs) are serious mental health conditions that predominantly emerge during adolescence and often persist into young adulthood (Treasure et al., 2020; Hammerle et al., 2016). They comprise a heterogenous group of clinical presentations but share the core feature of disturbed behaviours and/or perceptions towards either weight, body shape or eating. A recent study suggests that the global burden of disease (GBD) for EDs markedly increased from 1990 to 2021 among 10-24-year-olds (Liu et al., 2025). In this scenario, country-specific up to date prevalence estimates of EDs are essential to inform service planning, as well as resource allocation for existing services.

The epidemiological classification of eating disorders was significantly changed from DSM-IV to DSM-5. Previously, in DSM-IV, the eating disorders of clinical severity that did not meet full criteria for either anorexia nervosa or bulimia nervosa were categorised as Eating Disorder Not Otherwise Specified (EDNOS) (Fairburn & Bohn, 2005). DSM-5 aimed to reduce the number of EDNOS diagnosis (Ernst et al., 2017), by lowering the threshold for some eating disorders, for instance, removing the amenorrhea criterion from the Anorexia Nervosa diagnosis and introducing Other Specified Feeding or Eating Disorder (OSFED), a new diagnostic category, which comprises atypical Anorexia Nervosa, atypical Bulimia Nervosa, Purging Disorder and Night Eating Syndrome (*American Psychiatric Association*, 2013).

In practise, the overall effect of these changes in community adolescent samples seemed to be an increase in the estimated prevalence of any eating disorders, with an increased proportion of children and young people (CYP) assigned OSFED diagnoses. For example, in a community sample of German adolescents, prevalence estimates for any ED diagnoses increased from 0.48 (DSM IV) to 1.15% (DSM-5) while EDNOS/OSFED increased from 2.90 to 6.23% (Ernst et al., 2017) when applying DSM-IV and DSM-5 different diagnostic criteria, respectively. However, OSFED as a diagnostic category is notably absent from several epidemiological studies and the GBD Study only included estimates for anorexia and bulimia in both 2019 and 2021 (GBD 2021). A re-analysis of GBD data estimated that the study underestimated the prevalence of eating disorders by 41·9 million cases (95% uncertainty interval 27·9–59·0) by not including OSFED and binge eating disorders (Santomauro et al., 2021), underscoring the importance of robust populational studies with comprehensive coverage of all types of EDs.

DSM-5 also introduced a new eating disorder subtype, Avoidant and Restrictive Food Intake Disorder (ARFID). ARFID is marked by a persistent disturbance in feeding or eating which results in an inability to meet nutritional and/or energy requirements (*American Psychiatric Association*, 2013). Unlike anorexia nervosa, in ARFID food intake restriction can have different drivers, but it is not motivated by a desire for weight loss (Sader et al., 2023). Two recent surveillance studies of incident cases presenting to services in the United Kingdom and the Republic of Ireland (2.79 per 100 000) and Canada (2.02 per 100 000) reported similar incidence rate rates among children and young people (Sanchez-Cerezo et al., 2024; Katzman et al., 2021). However general population prevalence estimates of ARFID in CYP are still lacking. Recent Adult Psychiatric Morbidity Survey (APMS) reported an ED prevalence of 2.5 (0.8, 7.4) among 16-24 years old, however the survey authors acknowledge this as an underestimate (Ridout et al., 2025). Similarly, the psychossocial impairment and association with other mental health difficulties with EDs remains poorly characterised at populational level.

In this study, we use data from the nationally representative population-based survey (Mental Health of Children and Young People in England, MHCYP, 2023) to examine the epidemiology of Anorexia, Bulimia, Other Feeding or Eating Disorder (OSFED), Binge Eating Disorder and Avoidant Restrictive Food Intake Disorder (ARFID) among 11-25 year olds (Karimipour et al., 2026).

The study had two main aims. First, to establish the prevalence of Eating Disorders, by subtype, in a general population probability sample of children and young people. Second, to describe the psychosocial impairment associated with eating disorders, including self- and parent-reported psychological distress, self-harm and service use. We examine patterns of mental health difficulties associated with eating disorders diagnosis using validated measures such as the Strenghts and Difficulties Questionnaire (SDQ) and the General Health Questionaire (GHQ). We expected a positive association between ED and all indicators of impairment and service use.

## Methods

### Study sample (see Figure 1)

The MHCYP Survey 2023 included 2,370 children and young people (CYP) aged 8-25 years living in England (Karimipour et al., 2026), and was the fourth follow up of the 2017 MHCYP Survey. The initial 2017 survey included a stratified probability sample of 9,117 2-19 years olds who were living in England (Sadler et al., 2018). Participants were drawn from the NHS Patient Register using random probability sampling within 380 postcode sectors across England (Karimipour et al., 2026). The 2023 follow up sample frame comprised all those who agreed to re-contact. Participants were invited via a letter mailed to their homes with a link to an on-line survey. All participants in the MHCYP 2023 survey were screened for possible eating problems. Parents of 11-16 year olds, children aged 11-16 years and young people aged 17-25 years were asked to complete five screening questions taken from the Development and Well-Being Assessment (DAWBA) eating disorder module, and their responses were used to determine eligibility for a follow on study on EDs according to whether they screened positive (all invited, n=714 responses, n=563) or were achieved screen negatives (pre-selected random sample, n=547). The randomisation program aimed to achieve a 70:30 female:male gender ratio among screen negatives, because EDs are more common in young women, and a 50:50 split of young people (11-16 year olds) and emerging adults (17-25 year olds). The follow on study excluded 8-10 year olds (n=546), given extremely low numbers detected in our feasibility work. The 628 participants who screened negative and were not invited for the follow up study were retained as non cases, resulting in a final analytic sample of 1,738 (see Flowchart).

### Measures

#### Development and Well-Being Assessment (DAWBA)

The DAWBA is a multi-informant standardised diagnostic assessment with parent/carer and self-report versions for CYP aged 11 years and older (Goodman et al., 2000). Each module combines closed screening items, structured questions, and open-ended semi-structured probes that are transcribed verbatim.

In MHCYP 2023, the eating disorder module was completed by parents of 11-16-year-olds and by young people aged 11-25 years reporting on themselves. For young people aged 17 years and over self-report was the sole source. Of those invited to the second-stage assessment, 1,110 provided sufficient information for a DSM-5 clinical rating to be assigned. Most completed the module online, with telephone follow-up offered to non-responders. Eight six respondents were excluded as they did not sufficiently complete the DAWBA for their ED status to be determined.

Trained clinical raters (CF, AC, TF, IH, MK, OA, JO) reviewed parent and young person responses and assigned diagnoses according to DSM-5 criteria. Contentious or challenging diagnoses were reviewed with two senior consultant child and adolescent psychiatrists who specialised in EDs, and all diagnoses were reviewed by TF. Any eating disorder was defined as the presence of one or more of :anorexia nervosa, bulimia nervosa, binge-eating disorder, ARFID, and OSFED

#### Strengths and Difficulties Questionnaire (SDQ)

The SDQ is a 25-item behavioural screening questionnaire with parent- and self-report versions, widely validated in British general population samples (Goodman, 1997; Goodman, 2001). In MHCYP 2023, the parent-report version was completed by parents of 11-16-year-olds, and the self-report version by 11-25 year olds.

Items are rated on a three-point scale (not true, somewhat true, certainly true) and grouped into five subscales of five items each: emotional symptoms, conduct problems, hyperactivity/inattention, peer relationship problems, and prosocial behaviour. Scoring followed the official algorithm. Total difficulties was derived as the sum of the emotional symptoms, conduct problems, hyperactivity/inattention, and peer problems subscales (range 0-40). Higher scores indicate greater difficulty.

#### General Health Questionnaire (GHQ-12)

The GHQ-12 is a 12-item self-report screening measure of current psychological distress, developed for use in primary care and community settings (Goldberg & Williams, 1988). The GHQ-12 has been extensively validated in the UK population and displays good metric and scalar invariance across different ethnic groups (Anjara et al., 2020; King et al., 2023). Items are rated on a four-point scale with reference to the preceding two weeks. In MHCYP 2023 the GHQ-12 was completed by parents of 11-16-year-olds, reporting on their own mental health rather than that of their child, and by young people aged 17-25 years reporting on themselves.

Two scoring approaches are established for the GHQ-12: the binary method, which yields a caseness count, and the Likert method, which yields a continuous severity score ranging from 0-36. We used the Likert method because it retains information on symptom gradation (Anjara et al., 2020). Totals were calculated for participants completing all 12 items.

### Self-Harm

Self-harm was assessed with two items, using a four-week and a lifetime reference period respectively (parent version: “In the last 4 weeks, did your child try to harm themselves?” and “Over the whole of their lifetime, has your child ever tried to harm themselves?”). Parents reported for 11-16-year-olds and young people aged 17-25 years reported for themselves. Responses were coded as yes or no, and each item was modelled as a distinct outcome rather than combined, since the two reference periods capture different constructs.

The parent-reported four-week item is not reported. Under the Office of National Statistics (ONS) statistical disclosure control rules every reported cell must contain a minimum of 10 observations, and the number of events on this item among participants with an eating disorder fell below that threshold. Lifetime self-harm from both informants, and the young person-reported four-week item, all met the disclosure requirement and are reported.

### Special Educational Needs and Disabilities (SEND)

Parents of 11-16-year-olds were asked whether their child had special educational needs and/or disabilities (SEND). Responses were coded as present or absent; responses of “don’t know” and “prefer not to answer” were treated as absence of SEND, and those not applicable were excluded. SEND was available from parent report only and was therefore restricted to the 11-16 age band.

### Service and Medication Use

Service contact was assessed with a single multiple-response item: “Since [date one year previously], have you [or your child] been in contact with any of the following people or services because of worries about [your/their] mental health or emotional, behavioural or concentration problems?” Respondents could endorse any number of nine service categories: school-, college- or university-based mental health support; educational support services; primary health care; specialist mental health services (Child and Adolescent Mental Health Services, and Adult Mental Health Services in the young person version); physical health services; social care; private, paid-for services; Accident and Emergency; and a free-text “other” category. Contact with one or more of these was coded as a single binary indicator of any mental health service contact. Contact with a class teacher or member of college or university staff was recorded separately in the questionnaire and was not counted, as it does not constitute contact with a service.

Medication use was assessed with the item: “Since [date one year previously], have you [has your child] been prescribed any medication for a mental health or emotional, behavioural or concentration problem?” Responses were coded as yes or no. The same one-year reference period applied to both the parent and young person versions.

For both outcomes, parent report was used for 11-16-year-olds and young person self-report for 17-25-year-olds.

### Statistical Analyses

All analyses were conducted in R version 4.4.3 (R Core Team, 2025) using the survey package version 4.5 for design-based weighted estimation (Lumley, 2004).

Sociodemographic characteristics were compared between participants with and without any eating disorder using a survey-weighted t-test for age and Rao-Scott corrected chi-square tests for sex, ethnicity, and Index of Multiple Deprivation (IMD) quintile (see Table 1).

**Table 1.**
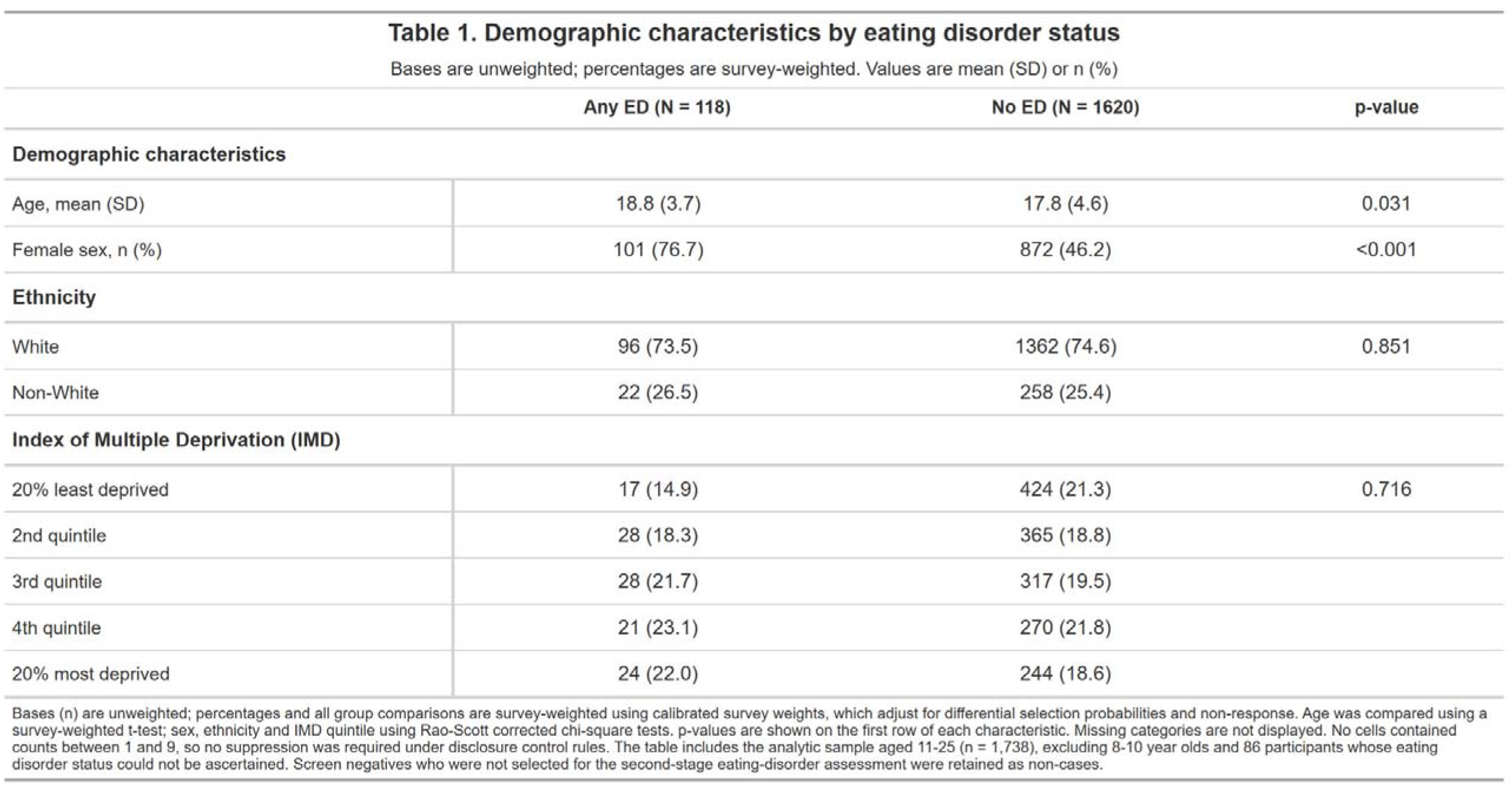
Demographic characteristics by eating disorder status.

Bases are unweighted; percentages are survey-weighted. Values are mean (SD) or n (%)
|  | Any ED (N = 118) | No ED (N = 1620) | p-value |
| --- | --- | --- | --- |
| **Demographic characteristics** |  |  |  |
| Age, mean (SD) | 18.8 (3.7) | 17.8 (4.6) | 0.031 |
| Female sex, n (%) | 101 (76.7) | 872 (46.2) | <0.001 |
| **Ethnicity** |  |  |  |
| White | 96 (73.5) | 1362 (74.6) | 0.851 |
| Non-White | 22 (26.5) | 258 (25.4) |  |
| **Index of Multiple Deprivation (IMD)** |  |  |  |
| 20% least deprived | 17 (14.9) | 424 (21.3) | 0.716 |
| 2nd quintile | 28 (18.3) | 365 (18.8) |  |
| 3rd quintile | 28 (21.7) | 317 (19.5) |  |
| 4th quintile | 21 (23.1) | 270 (21.8) |  |
| 20% most deprived | 24 (22.0) | 244 (18.6) |  |

Bases (n) are unweighted; percentages and all group comparisons are survey-weighted using calibrated survey weights, which adjust for differential selection probabilities and non-response. Age was compared using a survey-weighted t-test; sex, ethnicity and IMD quintile using Rao-Scott corrected chi-square tests. p-values are shown on the first row of each characteristic. Missing categories are not displayed. No cells contained counts between 1 and 9, so no suppression was required under disclosure control rules. The table includes the analytic sample aged 11-25 (n = 1,738), excluding 8-10 year olds and 86 participants whose eating disorder status could not be ascertained. Screen negatives who were not selected for the second-stage eating-disorder assessment were retained as non-cases.

Calibrated survey weights were applied to all analyses. These weights account for differential selection probabilities and non-response, supporting estimates representative of the population of England (Karimipour et al., 2026). Screen negatives who were not selected for the second-stage assessment were retained as non-cases, following the convention applied by NHS Digital to the corresponding derived eating disorder variables in this dataset. Weighted point prevalence was estimated overall, by sex, and by age band, with 95% confidence intervals (CI) calculated on the logit scale.

In all models, eating disorder status was the exposure and each mental health or impairment measure was the outcome. The outcomes analysed were SDQ total difficulties and GHQ-12 total score, treated as continuous, and SDQ predicted probable disorder, SEND, self-harm, medication use, and mental health service contact, treated as binary. Model family was determined by the distribution of the outcome: survey-weighted linear regression for continuous outcomes and survey-weighted logistic regression for binary outcomes. Separate models were fitted for parents, children (11-16 year olds) and young people (17-19 year olds), since instruments were administered to different groups and are not directly comparable across informants.

Adjustment for age (continuous) and sex was applied to all three SDQ total difficulties models, the young person-reported GHQ-12, both SDQ predicted probable disorder models, and the self-harm, medication use, service contact models and SEND support variable. This was because age and sex are established confounders for all these outcomes irrespective of who provides the report. The parent-reported GHQ-12 was not adjusted, as it measures the responding parent’s own psychological distress. Ethnicity and IMD quintile were not included as covariates, as neither showed evidence of association with eating disorder status (see Table 1).

The SDQ predicted probable disorder classification is generated by an algorithm drawing on three symptom domains. To establish whether the observed association was concentrated in a particular domain, we repeated the probable-disorder models using the corresponding domain-specific predicted diagnoses for emotional, conduct, and hyperactivity disorder again as binary probable versus unlikely or possible outcomes, rather than as continuous subscale scores; separately for each informant group and adjusted for age and sex (Supplementary Table S1).

All models used complete cases. Completion rates are reported in Appendix S1.

### Pre-registration

The analyses reported here were not pre-registered. This paper reports a secondary analysis of the Mental Health of Children and Young People in England (MHCYP) 2023 survey, commissioned by NHS Digital, for which the survey design, sampling strategy, questionnaire content and the two-stage procedure used to assess eating disorders were specified before data collection and are publicly documented in the published survey methodology and data quality statement (https://digital.nhs.uk/data-and-information/publications/statistical/mental-health-of-children-and-young-people-in-england/2023-wave-4-follow-up/data-quality-statement) and in the survey data resource profile (Karimipour et al., 2026).

**Figure 1.**
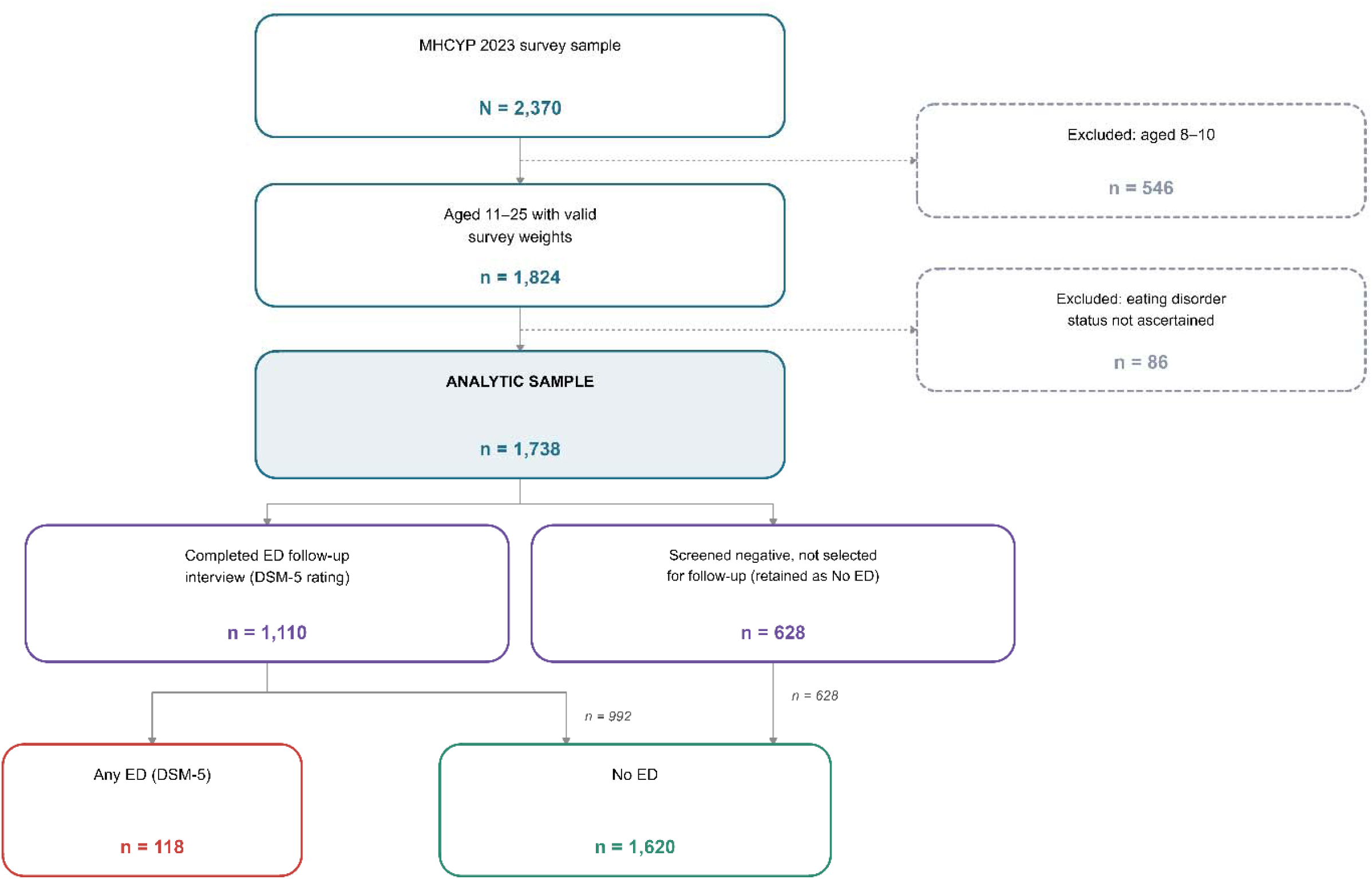
Flowchart

## Results

### Prevalence of Eating Disorders

The selected sample frame for the follow on study consisted of 1,277 individuals with responses for about 1,110 individuals (563 screen positives and 547 screen negatives), which represented a 87% response rate. In addition, 628 screen negatives not selected for the follow on study were retained as non cases, resulting in a final analytic sample of 1,738.

In weighted analyses, participants with an eating disorder were older than those without an eating disorder (mean age 18.8 versus 17.8 years), p=.031. Sex was also associated with eating disorder status, p<.001, with females comprising 76.7% of the eating disorder group compared with 46.2% of the group without an eating disorder. There was no evidence of differences between groups in ethnicity, p=.851, or IMD quintile, p=.716.

Weighted prevalence estimates for any eating disorder and by subtype are shown in Table 2. Any eating disorder affected 6.7% (95% CI 5.4–8.3) of 11-25-year-olds. Every subtype was more common in females, and the prevalence of any eating disorder was more than three times higher in females (10.7%, 95% CI 8.5–13.3) than in males (3.0%, 1.7–5.2). OSFED was by far the commonest diagnosis (3.5%, 2.6–4.7), accounting for approximately half of all cases, followed by bulimia nervosa (1.7%, 1.1–2.7) and anorexia nervosa (0.9%, 0.5–1.6). Binge eating disorder (0.6%, 0.3–1.3) and ARFID (0.2%, 0.1–0.6) were the least common.

**Table 2.**
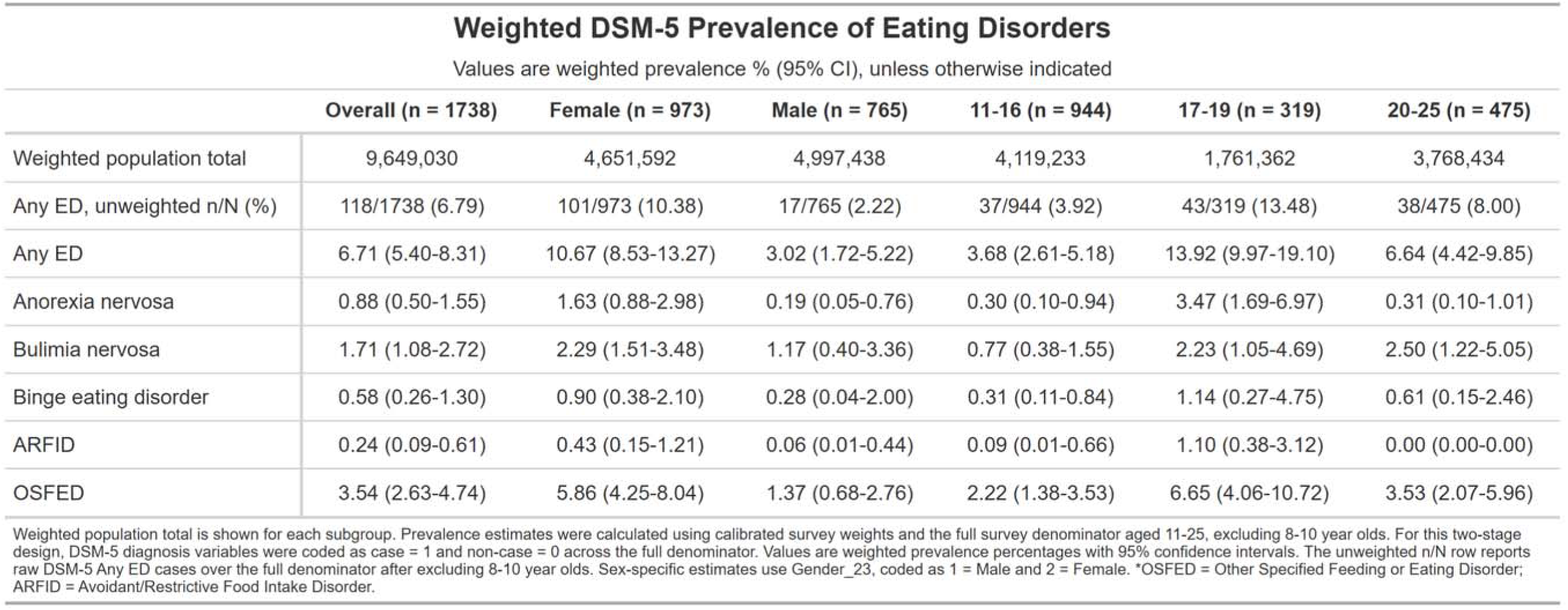
Weighted DSM-5 Prevalence of Eating Disorders.

| **Weighted DSM-5 Prevalence of Eating Disorders** |  |  |  |  |  |  |
| --- | --- | --- | --- | --- | --- | --- |
| Values are weighted prevalence % (95% CI), unless otherwise indicated |  |  |  |  |  |  |
|  | **Overall (n = 1738)** | **Female (n = 973)** | **Male (n = 765)** | **11-16 (n = 944)** | **17-19 (n = 319)** | **20-25 (n = 475)** |
| Weighted population total | 9,649,030 | 4,651,592 | 4,997,438 | 4,119,233 | 1,761,362 | 3,768,434 |
| Any ED, unweighted n/N (%) | 118/1738 (6.79) | 101/973 (10.38) | 17/765 (2.22) | 37/944 (3.92) | 43/319 (13.48) | 38/475 (8.00) |
| Any ED | 6.71 (5.40-8.31) | 10.67 (8.53-13.27) | 3.02 (1.72-5.22) | 3.68 (2.61-5.18) | 13.92 (9.97-19.10) | 6.64 (4.42-9.85) |
| Anorexia nervosa | 0.88 (0.50-1.55) | 1.63 (0.88-2.98) | 0.19 (0.05-0.76) | 0.30 (0.10-0.94) | 3.47 (1.69-6.97) | 0.31 (0.10-1.01) |
| Bulimia nervosa | 1.71 (1.08-2.72) | 2.29 (1.51-3.48) | 1.17 (0.40-3.36) | 0.77 (0.38-1.55) | 2.23 (1.05-4.69) | 2.50 (1.22-5.05) |
| Binge eating disorder | 0.58 (0.26-1.30) | 0.90 (0.38-2.10) | 0.28 (0.04-2.00) | 0.31 (0.11-0.84) | 1.14 (0.27-4.75) | 0.61 (0.15-2.46) |
| ARFID | 0.24 (0.09-0.61) | 0.43 (0.15-1.21) | 0.06 (0.01-0.44) | 0.09 (0.01-0.66) | 1.10 (0.38-3.12) | 0.00 (0.00-0.00) |
| OSFED | 3.54 (2.63-4.74) | 5.86 (4.25-8.04) | 1.37 (0.68-2.76) | 2.22 (1.38-3.53) | 6.65 (4.06-10.72) | 3.53 (2.07-5.96) |

Weighted population total is shown for each subgroup. Prevalence estimates were calculated using calibrated survey weights and the full survey denominator aged 11-25, excluding 8-10 year olds. For this two-stage design, DSM-5 diagnosis variables were coded as case = 1 and non-case = 0 across the full denominator. Values are weighted prevalence percentages with 95% confidence intervals. The unweighted n/N row reports raw DSM-5 Any ED cases over the full denominator after excluding 8-10 year olds. Sex-specific estimates use Gender\_23, coded as 1 = Male and 2 = Female. \*OSFED = Other Specified Feeding or Eating Disorder; ARFID = Avoidant/Restrictive Food Intake Disorder.

Prevalence varied markedly by age. Any eating disorder peaked among 17-19-year-olds (13.9%, 10.0–19.1), almost four times the estimate for 11-16-year-olds (3.7%, 2.6–5.2), with an intermediate estimate among 20-25-year-olds (6.6%, 4.4–9.9). This pattern was consistent across subtypes with the exception of bulimia nervosa, which was most prevalent in the oldest age band (2.5%, 1.2–5.1). No cases of ARFID were identified among 20-25-year-olds.

This is likely to reflect the rarity of the condition, but may also relate to ascertainment: eating disorder assessments above age 16 relied solely on self-report, with no parent informant available, and those most severely affected may be less likely to complete a self-report measure.

### Associated Impairment and Mental Health Difficulties

Associations between eating disorder diagnosis, self-harm, medication use and service contact are shown in Table 3. All estimates are adjusted for age and sex.

**Table 3.**
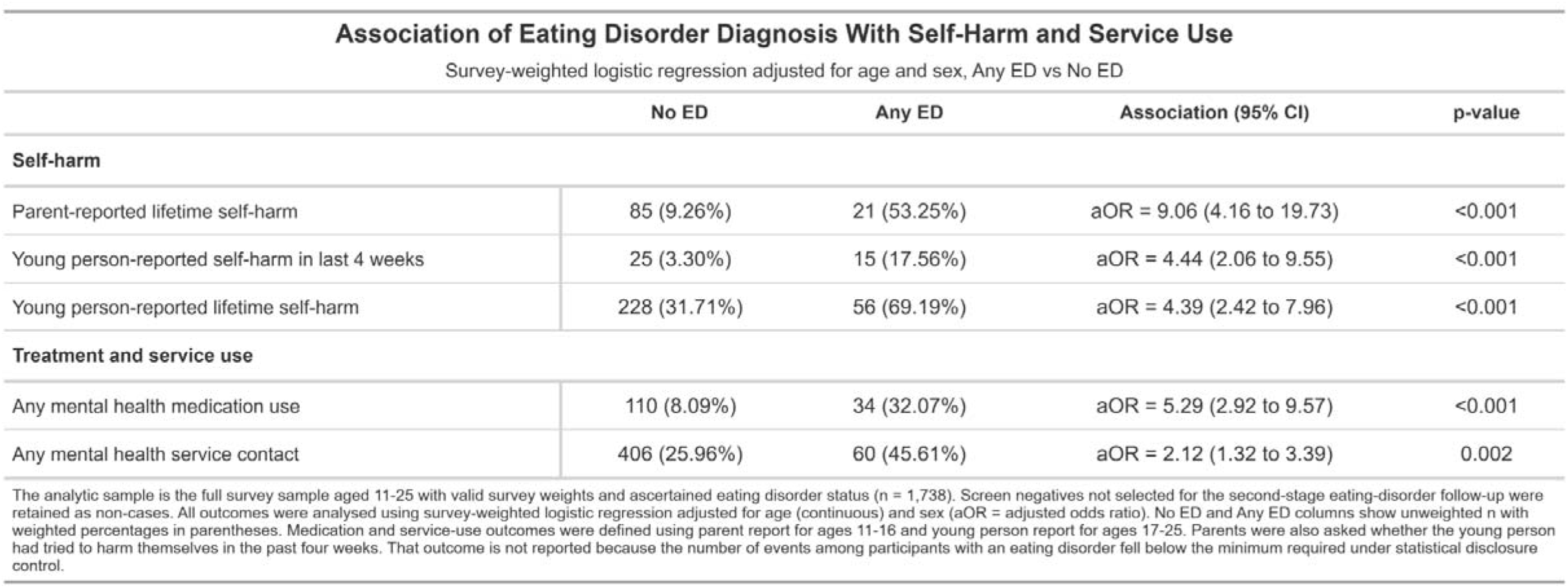
Association of Eating Disorder Diagnosis with Self-Harm and Service Use.

|  |  |  |  |  |
| --- | --- | --- | --- | --- |
| Survey-weighted logistic regression adjusted for age and sex, Any ED vs No ED |  |  |  |  |
|  | No ED | Any ED | Association (95% CI) | p-value |
| **Self-harm** |  |  |  |  |
| Parent-reported lifetime self-harm | 85 (9.26%) | 21 (53.25%) | aOR = 9.06 (4.16 to 19.73) | <0.001 |
| Young person-reported self-harm in last 4 weeks | 25 (3.30%) | 15 (17.56%) | aOR = 4.44 (2.06 to 9.55) | <0.001 |
| Young person-reported lifetime self-harm | 228 (31.71%) | 56 (69.19%) | aOR = 4.39 (2.42 to 7.96) | <0.001 |
| **Treatment and service use** |  |  |  |  |
| Any mental health medication use | 110 (8.09%) | 34 (32.07%) | aOR = 5.29 (2.92 to 9.57) | <0.001 |
| Any mental health service contact | 406 (25.96%) | 60 (45.61%) | aOR = 2.12 (1.32 to 3.39) | 0.002 |
| The analytic sample is the full survey sample aged 11-25 with valid survey weights and ascertained eating disorder status (n = 1,738). Screen negatives not selected for the second-stage eating-disorder follow-up were retained as non-cases. All outcomes were analysed using survey-weighted logistic regression adjusted for age (continuous) and sex (aOR = adjusted odds ratio). No ED and Any ED columns show unweighted n with weighted percentages in parentheses. Medication and service-use outcomes were defined using parent report for ages 11-16 and young person report for ages 17-25. Parents were also asked whether the young person had tried to harm themselves in the past four weeks. That outcome is not reported because the number of events among participants with an eating disorder fell below the minimum required under statistical disclosure control. |  |  |  |  |

Self-harm was substantially more common among CYP with an eating disorder. Among 11-16-year-olds, 53.3% of those with an eating disorder had ever self-harmed according to parent report, compared with 9.3% of those without (aOR=9.1, 4.2–19.7). Among 17--25-year-olds, 69.2% of those with an eating disorder reported having ever self-harmed, compared with 31.7% of those without (aOR=4.4, 2.4–8.0), and 17.6% reported self-harm within the preceding four weeks, compared with 3.3% of those without an eating disorder (aOR=4.4, 2.1–9.6).

Treatment and service contact were also more common. Mental health medication had been prescribed to 32.1% of CYP with an eating disorder compared with 8.1% of those without (aOR=5.3, 2.9–9.6), and 45.6% had been in contact with a mental health service in the preceding year compared with 26.0% (aOR=2.1, 1.3–3.4).

Associations with SEND, SDQ total difficulties, SDQ predicted diagnosis and GHQ-12 scores are shown in Table 4. There was no evidence of an association between eating disorder diagnosis and parent-reported SEND among 11-16-year-olds (24.3% versus 16.4%; OR=1.6, 0.7–3.6, p=.23).

**Table 4.** Association of Eating Disorder Diagnoses with Mental Health Difficulties.

| **Association of Eating Disorder Diagnosis With Special Educational Needs/Disabilities, SDQ Total Difficulties and Predicted Diagnosis, and GHQ-12 Scores** |  |  |  |  |
| --- | --- | --- | --- | --- |
| Survey-weighted models comparing Any ED vs No ED |  |  |  |  |
|  | No ED | Any ED | Association (95% CI) | p-value |
| **Parent-reported SEND (11-16)** |  |  |  |  |
| Special educational needs/disabilities | 143 (16.43%) | 10 (24.28%) | aOR = 2.12 (0.92 to 4.88) | 0.078 |
| **Parent-reported SDQ (11-16)** |  |  |  |  |
| Total difficulties | 9.64 (7.22) | 18.52 (8.64) | Adjusted Beta = 9.44 (6.45 to 12.42) | <0.001 |
| **Child-reported SDQ (11-16)** |  |  |  |  |
| Total difficulties | 10.46 (6.91) | 20.86 (5.69) | Adjusted Beta = 10.52 (8.12 to 12.93) | <0.001 |
| **Young person-reported SDQ (17-25)** |  |  |  |  |
| Total difficulties | 12.07 (6.03) | 17.03 (4.87) | Adjusted Beta = 4.39 (3.12 to 5.65) | <0.001 |
| **Parent-reported GHQ-12 (11-16)** |  |  |  |  |
| GHQ-12 total score | 12.06 (6.37) | 17.03 (7.68) | Beta = 4.97 (2.45 to 7.49) | <0.001 |
| **Young person-reported GHQ-12 (17-25)** |  |  |  |  |
| GHQ-12 total score | 12.11 (6.31) | 17.81 (6.57) | Adjusted Beta = 5.24 (3.28 to 7.20) | <0.001 |
| **Child+Parent-reported SDQ predicted diagnosis (11-16)** |  |  |  |  |
| Probable disorder (SDQ predicted diagnosis) | 178 (19.56%) | 27 (77.05%) | aOR = 14.72 (6.54 to 33.13) | <0.001 |
| **Young person-reported SDQ predicted diagnosis (17-25)** |  |  |  |  |
| Probable disorder (SDQ predicted diagnosis) | 140 (19.76%) | 43 (42.88%) | aOR = 2.54 (1.40 to 4.61) | 0.002 |

CYP with an eating disorder had markedly higher SDQ total difficulties scores, across every informant and both age bands. Among 11-16-year-olds, children with an eating disorder scored 10.5 points higher than their peers by self-report (20.9 versus 10.5; 8.1–12.9) and 9.4 points higher by parent report (18.5 versus 9.6; 6.5–12.4). Among 17-25-year-olds the difference was smaller but still substantial, at 4.4 points (17.0 versus 12.1; 3.1–5.7).

The same trend was evident using the SDQ predicted-diagnosis algorithm. Among 11-16-year-olds, 77.1% of those with an eating disorder were classified as probably having a psychiatric disorder, compared with 19.6% of those without (aOR=14.7, 6.5–33.1). Among 17-25-year-olds the corresponding proportions were 42.9% and 19.8% (aOR=2.5, 1.4–4.6).

Psychological distress was also elevated. Young people aged 17-25 with an eating disorder scored 5.2 points higher on the GHQ-12 than those without (17.8 versus 12.1; 3.3–7.2). Among 11-16-year-olds the GHQ-12 was completed by parents reporting on their own mental health rather than on their child’s; parents of children with an eating disorder scored 5.0 points higher than other parents (17.0 versus 12.1; 2.5–7.5).

## Discussion

We analysed the prevalence of DSM-5 eating disorders (EDs), and their associated mental health difficulties, impairment and treatment use, in a general population sample of CYP aged 11–25 in England. The prevalence of any ED was 6.7% (95% CI 5.4–8.3). In line with previous literature (Faria et al., 2026; Galmiche et al., 2019), we found EDs were more than three times more common in girls (10.7%) than boys (3.0%). We also found that the most prevalent type of ED, in both sexes, was OSFED (5.9% in girls and 1.4% in boys), replicating previous work (Santomauro et al., 2021; Solmi et al., 2016; Lindvall Dahlgren et al., 2017).

To our knowledge, this is the first European study to report general population prevalence of ARFID in CYP. We found a prevalence of 0.2% (0.1–0.6). A Taiwanese nationally representative study reported a similar 6-month prevalence of 0.3% (0.1–0.5) (Chen et al., 2019).

As hypothesised, we found an association between EDs and every form of impairment we measured. An ED diagnosis was associated with significantly higher SDQ total difficulties scores across all informants. The association was strongest for child self-report among 11-16-year-olds, whose mean total difficulties score was 20.9, some 10.5 points higher than their counterparts without EDs, compared with a difference of 4.4 points among 17-25-year-olds. The same age gradient was evident using the SDQ predicted-diagnosis algorithm: 77.1% of 11-16-year-olds with an ED were classified as probably having a psychiatric disorder, compared with 19.6% of those without (aOR=14.7, 6.5–33.1), whereas among 17-25-year-olds the corresponding figures were 42.9% and 19.8% (aOR=2.5, 1.4–4.6). Supplementary analyses by symptom domain indicated that this reflected broad, multi-domain difficulties in the younger group, whereas in older participants the association was largely confined to emotional symptoms. Notably, 69.2% of young people aged 17–25 with an ED reported having self-harmed in their lifetime, compared with 31.7% of those without an ED diagnosis, and 17.6% reported self-harm within the preceding four weeks compared with 3.3%. Among 11-16-year-olds self-harm was ascertained from parents rather than directly from the child, and the reported lifetime frequency was lower (53.3%). This partly reflects the younger age of this group, but it is also well established that parents typically underestimate self-injurious behaviour in their children (Breton et al., 2002; Krebs et al., 2025).

Together these findings suggest that early-onset EDs are accompanied by more extensive psychopathology and that younger adolescents may benefit from more intensive support. We found that 45.6% of those with EDs had been in contact with mental health services, compared with 26.0% of those without EDs; a UK-based community study across a wide age range (16–90) reported a similar rate of contact in the preceding year, at 30% (Solmi et al., 2016). However, more than half of young people meeting diagnostic criteria for an eating disorder had had no contact with any mental health service in the preceding year, and more than two thirds had received no medication; a substantial treatment gap in a group with demonstrably high levels of impairment.

Our findings have important policy implications. The finding that more than half of those meeting diagnostic criteria had no contact with mental health services in the preceding year suggests that current provision reaches only a minority of affected young people. To inform policy on an ongoing basis, and so that the burden of EDs can be appropriately quantified, continued data collection is needed. A series of nationally representative, general population surveys would allow us not only to quantify the prevalence and burden of EDs, but also to monitor ED trajectories in CYP.

At a clinical level, we replicated the finding that self-harm is common in CYP with EDs, extending it to a general population sample rather than the clinical samples used in most previous studies (Amiri & Khan, 2023; Moore et al., 2025; Meier et al., 2025). Routine enquiry about self-harm among young people presenting with disordered eating, in primary as well as specialist care, therefore appears warranted.

Strengths of our study include a representative population-based sample, multiple informants, and comprehensive diagnostic assessments by experienced clinicians following DSM-5 criteria. Our study also has important limitations. First, our analysis was based on the MHCYP 2023 data, which has a considerably smaller sample than its parent survey, MHCYP 2017 (Sadler et al., 2018; Karimipour et al., 2026). Attrition may have introduced bias, although we used calibrated weights to compensate for underrepresentation of certain groups. Second, the DAWBA screening items used in the first stage do not ask specifically about less typical symptoms of disordered eating, such as avoidant food behaviour. While the DAWBA has been validated in general population samples (Goodman et al., 2000), the absence of an ARFID specific screening instrument, together with the exclusion of 8-10-year-olds from the second stage, may have led to an underestimate of ARFID prevalence. Finally, small numbers of ED cases within individual ethnic groups meant that, for disclosure reasons, we were unable to report prevalence by ethnic group.

In summary, we found that EDs are common and that they are associated with high levels of impairment, self-harm, and comorbid mental health difficulties, particularly among 11-16-year-olds. Despite this, most affected young people had no recent contact with mental health services. Further research should focus on improving recruitment of ethnic minority groups, whose experiences remain underrepresented in current studies, and on developing screening measures that reflect the heterogeneity of ED presentations. Future work should also explore whether impairment predicts eating disorder trajectories and prognosis, particularly given the high levels of impairment we observed among 11-16-year-olds.

## Ethics statement

The MHCYP 2020 follow-up survey was reviewed and received ethical approval from the National Statistician’s Data Ethics Advisory Committee in May 2020. This approval included future follow-up surveys. All procedures contributing to this work comply with the ethical standards of the relevant national and institutional committees on human experimentation and with the Helsinki Declaration of 1975, as revised in 2008. This secondary data analysis received approval from the UK Data Service and from the data controllers under project 273302 on January 2025.

Informed consent was obtained at each survey wave. Participants who agreed at interview to be recontacted for future research, and who continued to agree at each wave in which they subsequently took part, were invited to the 2023 survey; consent to recontact was held for 6,898 children and young people aged 8 to 25 years. In 2022 and 2023, children aged 16 years and their parents were asked the consent questions on data linkage and future research, whereas in 2017, 2020 and 2021 these were asked of parents only.

## AI statement

The authors acknowledge the use of Claude Opus (Anthropic), accessed between August and September 2026. Claude was used to check grammar and spelling; to reduce the manuscript word count; to format the reference list to journal requirements by condensing and adapting content already present in the manuscript, without generating any new content. All AI-generated suggestions and outputs were reviewed, revised and approved by the authors, who take full responsibility for the accuracy and integrity of the work.

## Key points

What is known: DSM-5 broadened eating disorder classification, but general population prevalence estimates using DSM-5 criteria in children and young people are scarce, and population-level estimates for ARFID are lacking.

What is new: In a nationally representative English sample of 11-25-year-olds, 6.7% met DSM-5 criteria for an eating disorder, peaking at 13.9% among 17-19-year-olds, and OSFED accounted for approximately half of all cases.

This is the first European general population estimate of ARFID prevalence in this age group (0.2%).

What is relevant: Eating disorders were associated with markedly elevated self-harm, psychological distress and comorbid difficulties, particularly among 11-16-year-olds.

More than half of affected young people had no mental health service contact in the preceding year, indicating a substantial treatment gap relevant to service planning and commissioning.

## Acknowledgements

We would like to acknowledge NatCen and the Office for National Statistics for their logistical support in running the MHCYP surveys.

## Data availability statement

The data supporting the findings of this study are available from the UK Data Service. The Mental Health of Children and Young People in England 2023 survey dataset is available to researchers on application, under study number SN 9476, subject to the UK Data Service’s access conditions. No new data were generated by the present analysis.

