## Supplementary Information for "Prevalence of DSM-5 eating disorders and associated impairment and service use among children and young people in England: analysis of a general population survey"

Appendix S1 – Completion rates for measures

The completion rate was 100% for the parent-reported SDQ, the young person-reported SDQ, SEND, and both SDQ predicted-diagnosis classifications; 99.3% for the parent-reported GHQ-12 (937/944) and 98.0% for the young person-reported GHQ-12 (778/794). Completion was similarly high for the impairment outcomes: 99.8% for parent-reported lifetime self-harm (942/944), 98.7% and 98.9% respectively for young person-reported four-week (784/794) and lifetime (785/794) self-harm, 98.9% for medication use (1,719/1,738), and 99.0% for mental health service contact (1,721/1,738). Completion was lower for the child self-reported SDQ, at 65.5% of eligible 11-16-year-olds (618/944). Missingness rates were similar in both groups: this measure was completed by 65.3% (592/907) of participants without an ED and by 70.3% (26/37) of those with an eating disorder. Completion was instead patterned primarily by age, rising from 56% at age 11 to 73% at age 16.

In regression models, complete-case analysis remains unbiased when the likelihood of providing data depends on variables included in the model, but can be biased when it depends on the outcome itself. Non-completion here was strongly patterned by age, which is included as a covariate, and differed little by eating disorder status. The relevant question is therefore whether non-completion was also related to the child's own level of difficulty. This cannot be tested directly, since the self-report score is unobserved for those children. The parent-reported SDQ, however, was available for all 944 eligible 11- to 16-year-olds, including the 326 who did not complete the self-report, and provides an indirect check. Children who did not complete the self-report had slightly higher parent-reported total difficulties than those who did (mean 10.43 versus 9.44; difference 0.99, 95% CI −0.04 to 2.02, p = .06), suggesting any residual bias is small.

Table S1 – sensitivity analysis – SDQ probable disorder

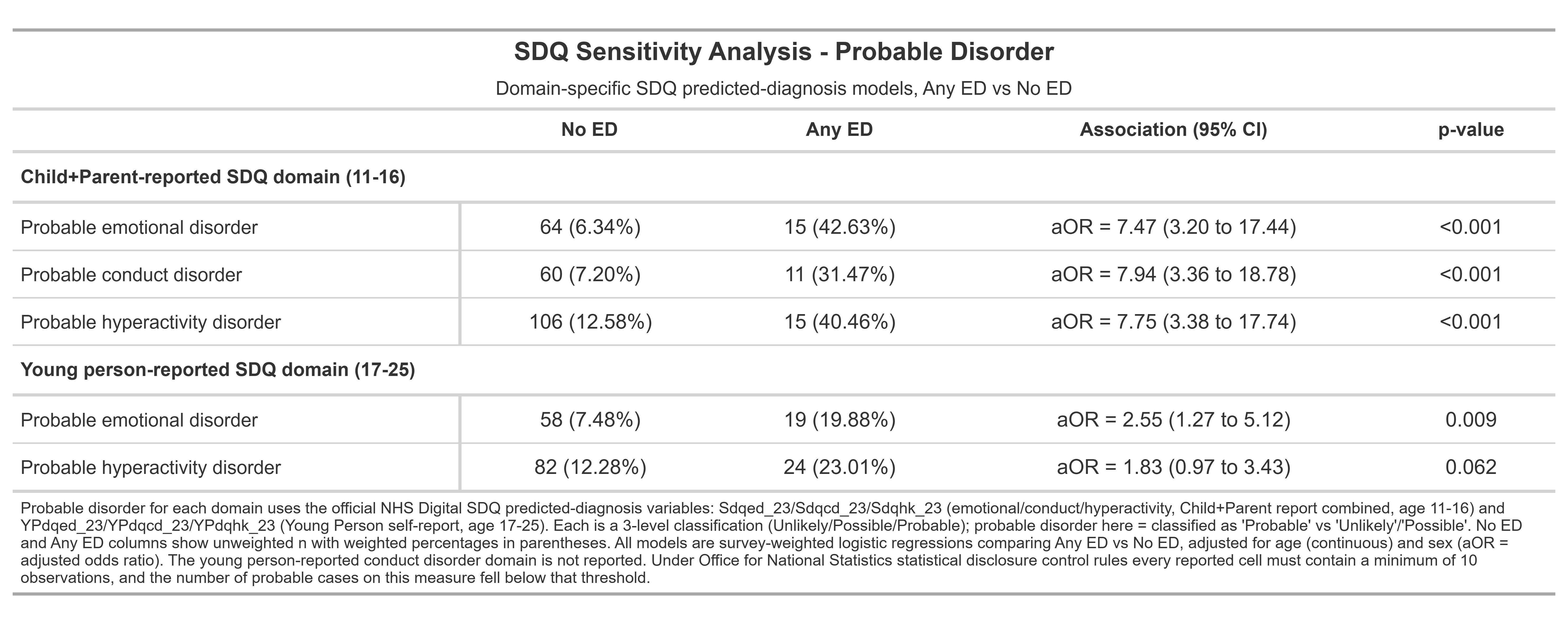
